# Circulating Adenoid Cystic Carcinoma associated MYB transcripts enable rapid and sensitive detection of metastatic disease in blood liquid biopsies

**DOI:** 10.1101/2024.10.15.24315549

**Authors:** Acadia H.M. Moeyersoms, Kendall W. Knechtel, Andrew J. Rong, Ryan A. Gallo, Michelle Zhang, Harper M. Marsh, Zoukaa B. Sargi, Jason M. Leibowitz, Francisco J. Civantos, Donald T. Weed, Sander R. Dubovy, David T. Tse, Daniel Pelaez

## Abstract

Adenoid cystic carcinoma (ACC) is a rare and lethal malignancy that originates in secretory glands of the head and neck. A prominent molecular feature of ACC is the overexpression of the proto-oncogene MYB. ACC has a poor long-term survival due to its high propensity for recurrence and protracted metastasis. Currently, clinical technologies lack the efficiency to distinguish patient prognosis prior to its redevelopment. We hypothesize that metastatic ACC can be detected by monitoring tumor-specific MYB expression in patients’ blood. We developed a quantitative polymerase chain reaction (qPCR) assay for MYB transcripts and screened blood samples from four patient cohorts: no history or evidence of ACC (n=23), past history of ACC and no evidence of disease (NED) for greater than three years (n=15), local ACC (n=6), and metastatic ACC (n=5). Our assay detected significantly elevated levels of MYB transcripts in the metastatic ACC cohort (p < 0.01). Receiver operating characteristic (ROC) curves comparing metastatic to NED and metastatic to local disease were significant, with p values < 0.0001 and 0.0008, respectively. Single-cell RNA sequencing (scRNA-seq) of blood from metastatic ACC identified a cluster of circulating tumor cells (CTCs) expressing MYB. Here, we report a sensitive, cost-effective, and minimally invasive diagnostic test that leverages tumor-specific signatures to screen for metastatic ACC disease, potentially enhancing detection earlier than the current clinical standard.

## 1 Background

Adenoid cystic carcinoma (ACC) is a rare but lethal epithelial cancer that predominantly arises from the secretory glands of the head and neck (1, 2). Despite surgical excision followed by radiation, the prognosis for patients with ACC remains poor due to high rates of recurrence and metastasis, with the 5, 10, and 20-year survival rates reported as 68-80%, 60%, and 28% respectively (3-5). Current surveillance methods involve periodic scans; however, technical complexities, limited access, and financial constraints contribute to patient attrition.

Here, we evaluate whether the known molecular signatures that give rise to ACC tumors could be harnessed for the development of a blood-based molecular assay to monitor patients with ACC. The most prominent molecular feature of ACC tumors is the overexpression of the MYB proto-oncogene, which is often translocated with other genomic loci to establish a feedforward mechanism that amplifies its expression in ACC (Figure 1A-B) (6-9). The reliance of ACC tumors on MYB expression to drive its survival and proliferation allows us to evaluate whether qPCR probes mapping to the 5’ regulatory elements of MYB can detect and discern metastatic disease from blood of patients with ACC from those with non-disseminated tumors and other, non-cancerous sources of MYB expression.

**Figure 1:**
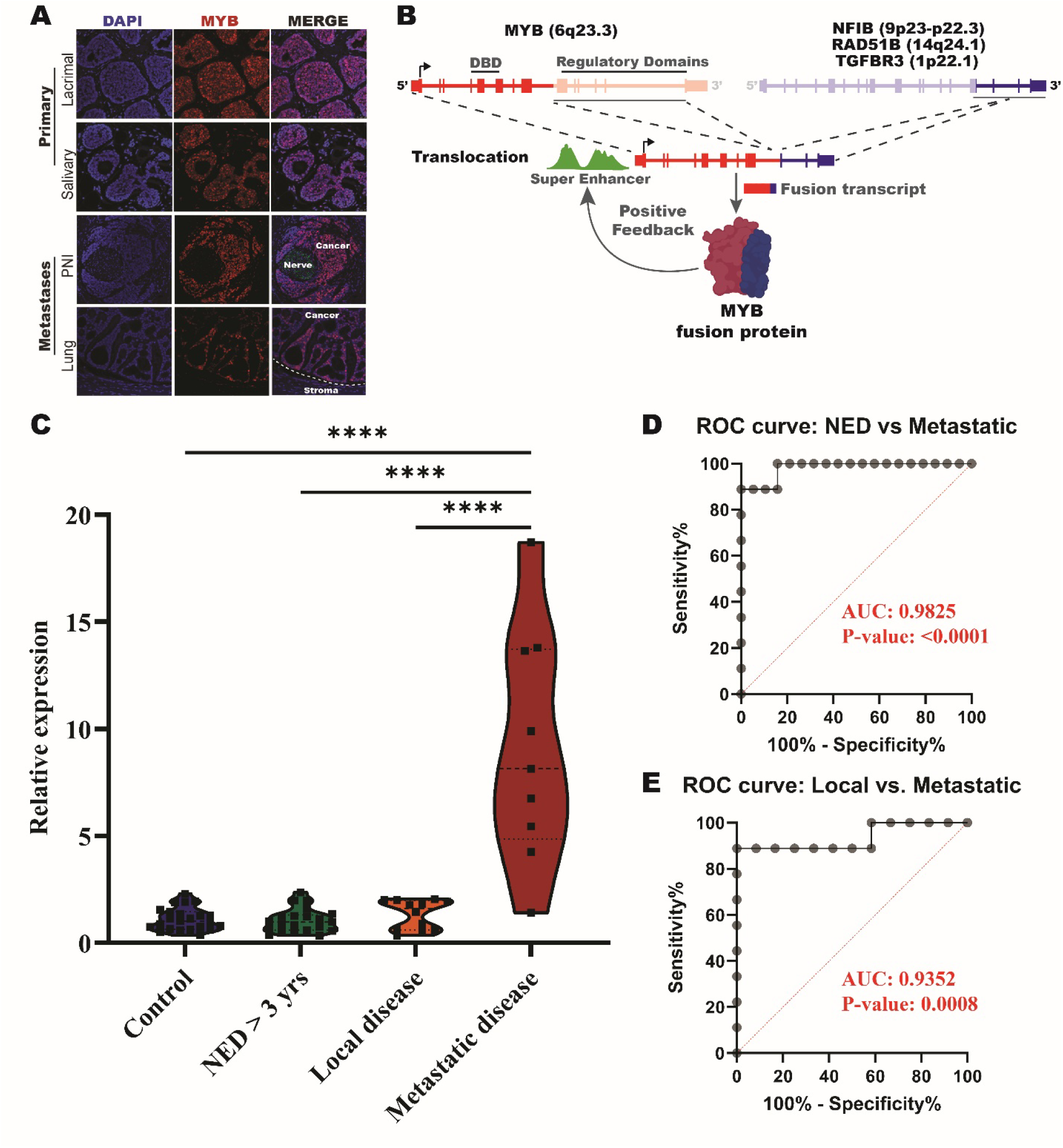
Detection of MYB in whole blood for patients with metastatic ACC. (**A**) Immunofluorescent imaging of MYB expression in primary and metastatic tumor samples. (**B**) Schematic of MYB translocation events that occur with NFIB, RAD51B, and TGFBR3. (**C**) Violin plot of relative expression of MYB primer pair spanning exons 2-3 in control, NED, local, and metastatic patients. ^****^ = p-value < 0.0001. (**D**) ROC curve of NED > 3 years verse metastatic results of whole blood MYB relative expression test. (**E**) ROC curve of local disease verse metastatic results of whole blood MYB relative expression test.

## 2 Methods

See supplemental information for detailed methods.

### 2.1 Peripheral blood sample collection and processing

The University of Miami Institutional Review Board approved this study (IRB 20090524 and 20210471). The study was conducted in a Health Insurance Portability and Accountability Act of 1996–compliant manner. Blood samples were collected from individuals with no history or evidence of ACC (n=23), past history of ACC and no evidence of disease (NED) (n=15), local ACC (n=6), and metastatic ACC (n=5) (Table 1) and frozen at -80°C until processed. Blood RNA was extracted and reverse-transcribed to cDNA. RT-qPCR was conducted and relative expression of MYB was quantified using the geometric mean of the Ct values for the two housekeeping genes and MYB amplification in the control samples (Supplementary Table 1) (10).

**Table 1:** Clinical demographic table.

|  | Sample<br>(n = 25) |
| --- | --- |
| **Patient demographics** |  |
| Age at diagnosis | 46.4 (SD = 13.6, Range: 20 – 71) |
| Gender |  |
| Male | 16 |
| Female | 9 |
| Affected Side |  |
| Right | 14 |
| Left | 9 |
| Not sided | 2 |
| **Tumor characteristics** |  |
| ACC location |  |
| Salivary | 2 |
| Sinus | 4 |
| Oropharyngeal | 1 |
| Larynx | 1 |
| Lacrimal | 17 |
| ACC subtype |  |
| **Crib** |  |
| **Solid variant** | 6 |
| **Crib/Solid** | 2 |
| **Crib/Basa** | 1 |
| **Crib/Tub** | 1 |
| **Unavailable** | 4 |
|  | 11 |
| **Bone involvement** |  |
| **T stage on presentation** |  |
| **1** | 5 |
| **2** | 5 |
| **3** | 3 |
| **4** | 10 |
| **Unavailable** | 3 |
| **N stage on presentation** |  |
| **0** | 21 |
| **3** | 1 |
| **Unavailable** | 3 |
| **M stage on presentation** |  |
| **0** | 22 |
| **1** | 0 |
| **Unavailable** | 3 |
| **Current clinical status** |  |
| **Post treatment local recurrence/metastasis** |  |
| **Local** | 6 |
| **Met (total)** | 5 |
| **Deceased** | 3 |
| **NED** | 15 |

### 2.2 Preparation of peripheral blood mononuclear cell (PBMC) fraction for single-cell RNA sequencing (scRNA-seq)

PBMC fractions were isolated by centrifugation. scRNA-seq library was prepared using the 10X Genomics Chromium Single Cell 5’ v2.0 chemistry (10x Genomics, Pleasanton, CA) according to manufacturer’s instructions. We used the CellRanger pipeline to process and align to the GRCh38 human genome reference. Further analysis was computed using various programs and references.

## 3 Results

### 3.1 Probes spanning the exon 2-3 boundary of MYB discern metastatic blood samples from other cohorts

We designed qPCR primer pairs spanning exon-exon boundaries throughout the 5’ end of the MYB gene body and validated all probes against a primary ACC tumor (Supplementary Figure 1). Subsequently, we tested six of these MYB probes in blood from patients with metastatic ACC. Only the primer pair spanning the exon 2 to 3 boundary detected an elevated MYB signal (Supplementary Figure 2). Utilizing this primer pair, we observed a significantly elevated level of MYB expression (p < 0.0001) in whole blood from our patients with metastatic ACC. In contrast, no significant level of MYB expression in blood was detected among the control, NED, or local disease cohorts (Figure 1C and Supplementary Figure 3). Using primer pairs that amplify products between exon 14 and 15 of MYB, we further validated that the elevated MYB signal detected in metastatic ACC blood originates from the enriched 5’ MYB transcription (Supplementary Figure 4).

ROC curves for metastatic versus NED and metastatic versus local disease were significant with p values of <0.0001 and 0.0008, respectively. The area under the curve (AUC) for both ROC curves were 0.9825 and 0.9352, respectively, indicating a highly significant and sensitive test (Figure 1C and D).

### 3.2 Circulating tumor cells (CTCs) are the source of elevated MYB in metastatic ACC blood

To elucidate the source of the elevated MYB signal detected in metastatic ACC blood samples, we isolated RNA from the plasma and mononuclear cell (PBMC) fractions to determine if the MYB signal arose from cell-free (plasma) or cell based (PBMCs) RNA sources. Our analysis revealed that the MYB level in the PBMC layer was 258.5-fold higher than that of the plasma (Figure 2A). Expanding on this, we performed scRNA-seq on the PBMC fraction of metastatic ACC blood (T6). The sequencing run captured 4,852 cells. Immune cell clusters were identified based on gene expression, whereas one cluster presented with the highest expression of MYB and did not have any immune cell gene expression signatures (indicated as cluster of interest in UMAP (n = 18 cells)) (Figure 2B). Upon integration of the scRNA-seq dataset with our previously reported spatial transcriptomic profiling of ACC (11), this cluster of interest was determined to have significant overlap in several ACC-specific markers, indicating that this cluster represents a circulating tumor cell (CTC) population (Figure 2C-E and Supplementary Figure 5).

**Figure 2:**
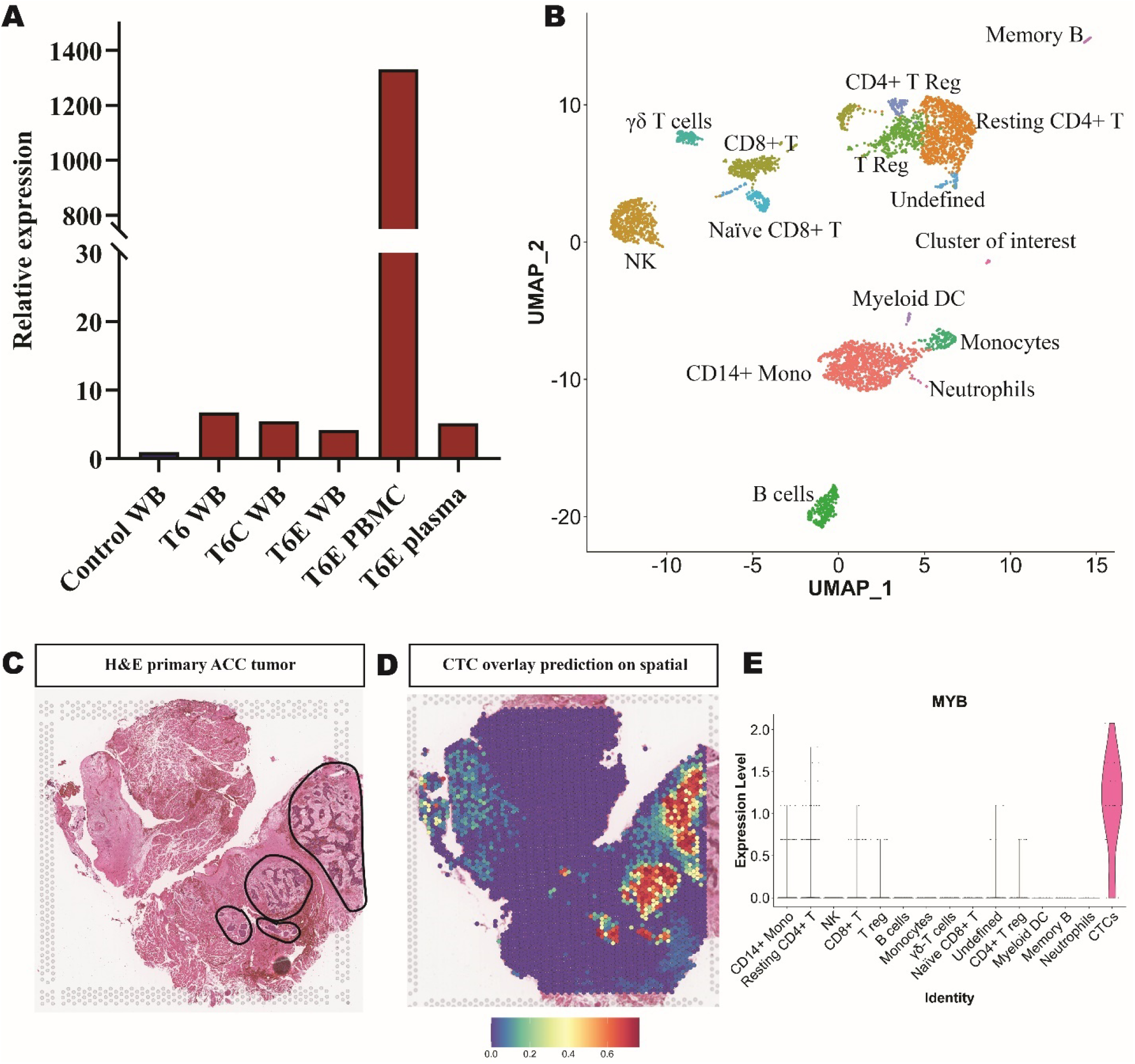
Circulating tumor cell identification in PBMCs of metastatic ACC patient. (**A**) Tracking patient T6 MYB relative expression over time in whole blood (WB), PMBC, and plasma samples. (**B**) UMAP of 15 clusters from sample T6 PBMC single-cell sequencing experiment with cell type identifications. (**C**) Hematoxylin and eosin (H&E) image of primary ACC tumor used for spatial transcriptomics with circled tumor foci indicated. (**D**) Probability prediction of single-cell cluster of interest cell type overlaid on spatial transcriptomic primary ACC tumor sample. (**E**) Violin plot of MYB expression in PBMC single-cell sequencing.

## 4 Discussion

We have developed a liquid biopsy that can detect a cancer specific signature from blood samples of patients with metastatic ACC. Our RT-qPCR assay detects tumor-specific MYB expression specifically and sensitively in a rapid and cost-effective manner. We show that the relative expression of MYB significantly differentiates metastatic ACC blood samples from samples collected from controls, NED, and local disease patients. Longitudinal follow-up revealed elevated MYB signal in one patient (T5) who was diagnosed with metastatic ACC by imaging 11-months after our first detection of elevated MYB in their blood sample. Subsequent blood draws from this patient (T5B, T5C) after their clinical metastatic diagnosis show progressively higher levels of MYB expression, commensurate with their disease burden (Supplementary Figure 6). About 10% ACCs do not exhibit MYB overexpression, which would require initial screening for MYB expression in primary tumor before employing out assay for surveillance. Yet, for the vast majority of patients with ACC, our assay may decrease unnecessary imaging, enable earlier interventions in the setting of protracted metastasis, and ultimately improve long-term prognosis.

In interrogating the source of the MYB signal, we discovered a CTC population in metastatic ACC blood (Figure 2C-D and Supplementary Figure 5). To our knowledge, only one previous report of CTCs was identified in ACC patients’ blood, which were isolated using the general marker pan-cytokeratin (12). In our analysis, keratin 8 had minor preferential expression in the ACC CTC cluster (Supplementary Figure 7). Transcriptomic analysis revealed high Sox4 expression in ACC CTCs, which may be a potential additional biomarker. However, qPCR probing of Sox4 expression lacked the diagnostic power provided by MYB (Supplementary Figure 8).

We acknowledge that a limitation of the current findings is the size of our sample cohorts, partly due to the rarity and lethality of ACC itself. A randomized prospective clinical study is warranted to ascertain the true diagnostic power and sensitivity of this assay to discern regional lymph node metastases in addition to distal metastases. We are currently working on establishing the protocols for such as study at our institution.

## 5 Conclusions

The findings of this study provide encouraging evidence that leveraging the key signature of ACC can facilitate the development of a simple, sensitive, and cost-effective metastatic screening tool for ACC management. Upon clinical validation, we envision a test that could be implemented for continuous monitoring of MYB levels in patients with a history of ACC, thereby enabling the stratification of individuals who may require confirmatory imaging or more frequent screenings.

## Supporting information

Supplemental Figures mentioned throughout the manuscript

Supplemental Methods, giving more in depth information on the material

## Data Availability

All data produced in the study is available upon request to the corresponding author.

## Acknowledgements

We would like to thank all participants who donated blood to this study. We would also like to thank University of Miami Miller School of Medicine Onco-Genomics Sequencing Core.

## Abbreviations

ACC: Adenoid cystic carcinoma
AUC: Area under the curve
CTC: Circulating tumor cells
NED: No evidence of disease
PBMC: Peripheral blood mononuclear cells
ROC curve: Receiver operating characteristic curve

## Declarations

### Ethics approval and consent to participate

The University of Miami Institutional Review Board approved this study (IRB 20090524 and 20210471). All subjects gave a written informed consent to participate in the study. The study was conducted in a Health Insurance Portability and Accountability Act of 1996–compliant manner.

### Consent for publication

All authors give consent for the publication of the following content.

### Availability for data and materials

Data presented is all available upon request to the corresponding author.

### Competing interests

The authors declare that there are no competing interests.

### Funding

Research reported in this publication was supported in part by the Dr. Nasser Al-Rashid Orbital Vision Research Endowment 700975 (D.T.T. and D.P.), a Department of Defense (DOD) Rare Cancers Research Program Grant W81XWH2211079 (D.P.), and the Adenoid Cystic Carcinoma Research Foundation (ACCRF) AWD006393 (D.P.). The Bascom Palmer Eye Institute is supported by NIH Center Core Grant P30EY014801 and a Research to Prevent Blindness Unrestricted Grant GRoo4596 (New York, NY, USA). The Sylvester Comprehensive Cancer Center shared resources are supported by the NCI Core Grant P30CA240139. The content is solely the responsibility of the authors and does not necessarily represent the official views of the NIH.

### Authors’ contributions

A.H.M.M, R.A.G, H.M.M, Z.B.S, J.M.L, F.J.C, D.T.W, and D.P were involved with the methodology, administration, and project resources. A.H.M.M, M.Z, and S.R.D conducted data analysis and interpretations. A.H.M.M, K.W.K, R.A.G, and D.P drafted and revised the manuscript. D.T.T and D.P provided financial support and guidance. All authors approved of the final manuscript.

## Figure legends

**Supplementary Figure 1: Amplification plot of preliminary MYB primer screening on primary ACC tumor tissue**. Quant Studio 7 amplification plot output of 10 MYB primers with cDNA from ACC tumor tissue.

**Supplementary Figure 2: Primer screen on blood samples for MYB amplification in metastatic samples**. Six primer pairs spanning exons 2-8 run on control and ACC patient blood to identify the difference between metastatic samples compared to disease free and control patients.

**Supplementary Figure 3: Relative expression of individual patients for MYB primer pair amplification of exons 2-3**. Relative expression levels of control, NED, local, and metastatic MYB whole blood levels.

**Supplementary Figure 4: Relative expression of MYB exons 14-15 in patient whole blood**. Endogenous levels of MYB in whole blood measured by the amplification of MYB exons typically lost in translocation events.

**Supplementary Figure 5: Violin plots of 30 genes overexpressed in spatial transcriptomic primary ACC tumor and circulating tumor cell cluster from blood scRNA-seq**. Violin plots of genes that are differentially expressed in both primary tumor and CTCs in blood.

**Supplementary Figure 6: Progression of MYB level in sample T5 overtime**. Relative expression of MYB in whole blood for sample T5 (February 2020), T5B (April 2021), and T5C (December 2021).

**Supplementary Figure 7: Keratin expression in different clusters from blood single-cell sequencing on metastatic ACC patient**. RNA expression for Keratins 1-30 were examined and detection of RNA for keratin 1, 2, 5, 7, 8, 10, 17, 18, and 23 was observed.

**Supplementary Figure 8: Examining Sox4 expression in whole blood via RT-qPCR**. Relative expression of Sox4 for 4 different primer pairs amplifying varying regions of the gene for control, NED, local, and metastatic disease patients.

**Supplementary Table 1:**
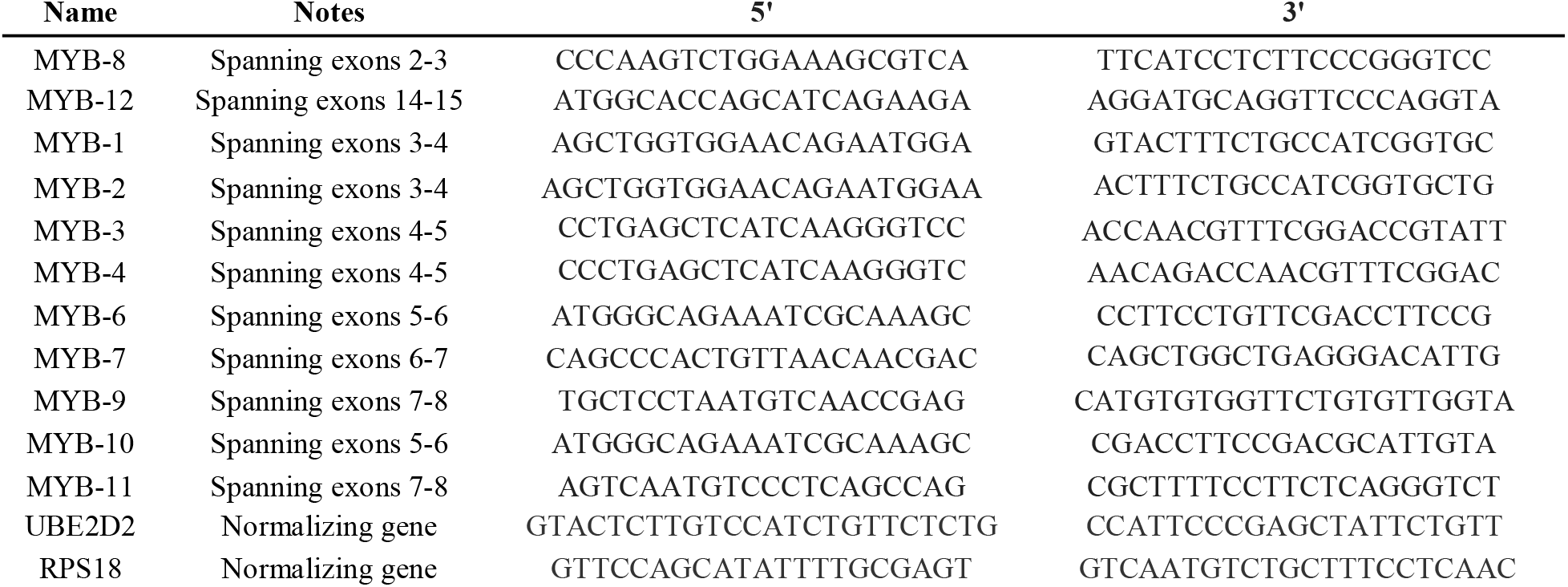
Table of primers used in RT-qPCR experiments.

## Notes

### Competing Interest Statement

The authors have declared no competing interest.

### Author Declarations

The Institutional Review Board of The University of Miami gave ethical approval for this study (IRB 20090524 and 20210471).

