## Supplemental Figures mentioned throughout the manuscript for "Circulating Adenoid Cystic Carcinoma associated MYB transcripts enable rapid and sensitive detection of metastatic disease in blood liquid biopsies"

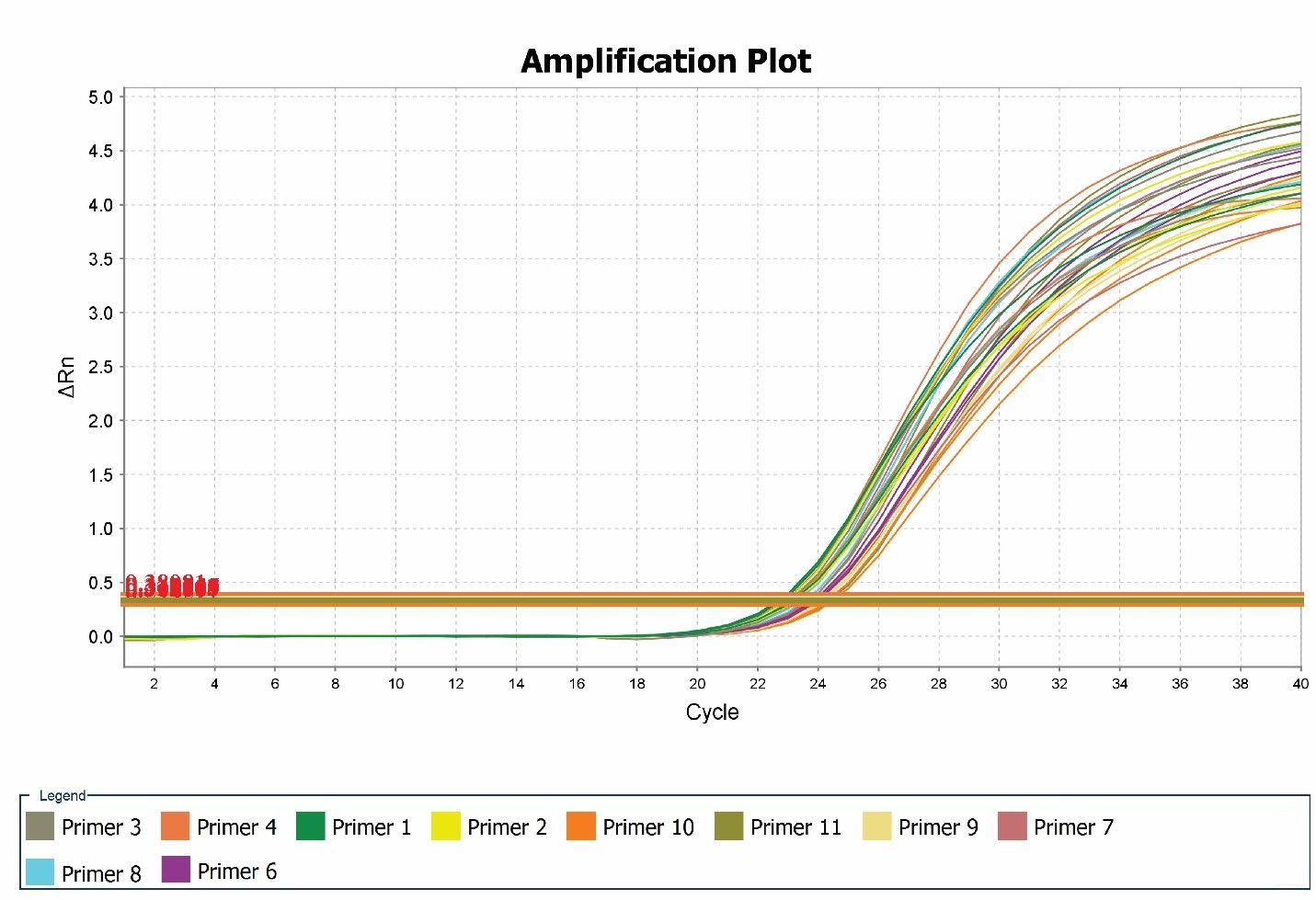

**Supplementary Figure 1.**

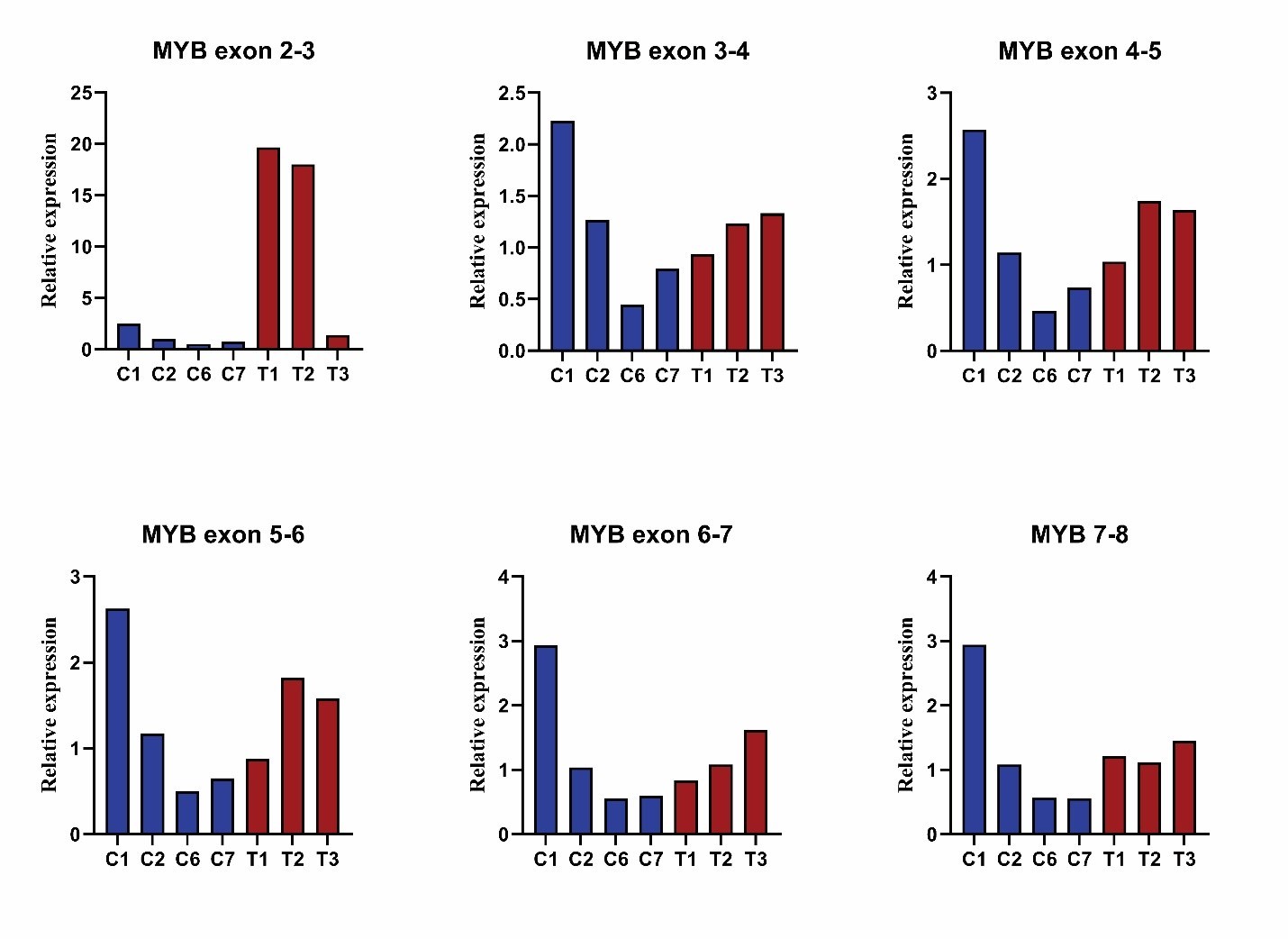

**Supplementary Figure 2.**

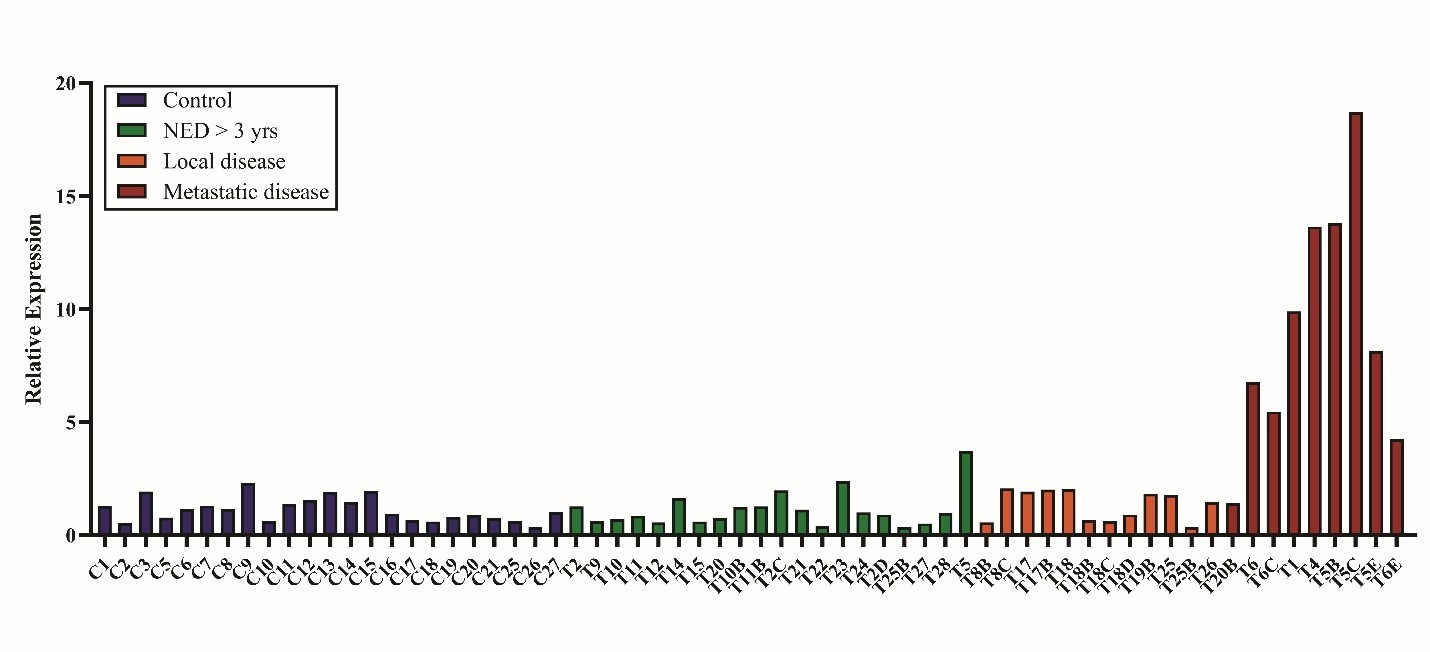

**Supplementary Figure 3.**

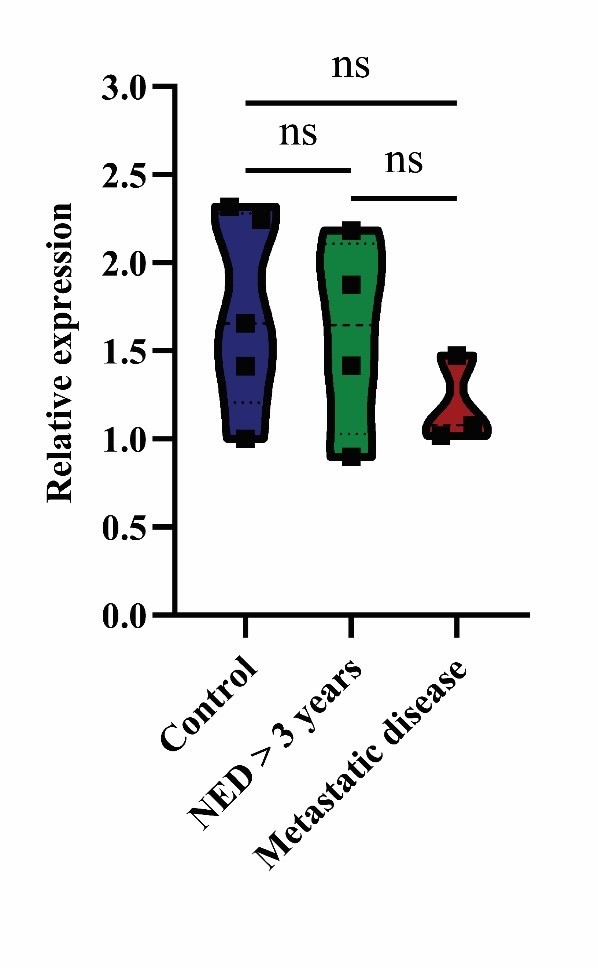

**Supplementary Figure 4.**

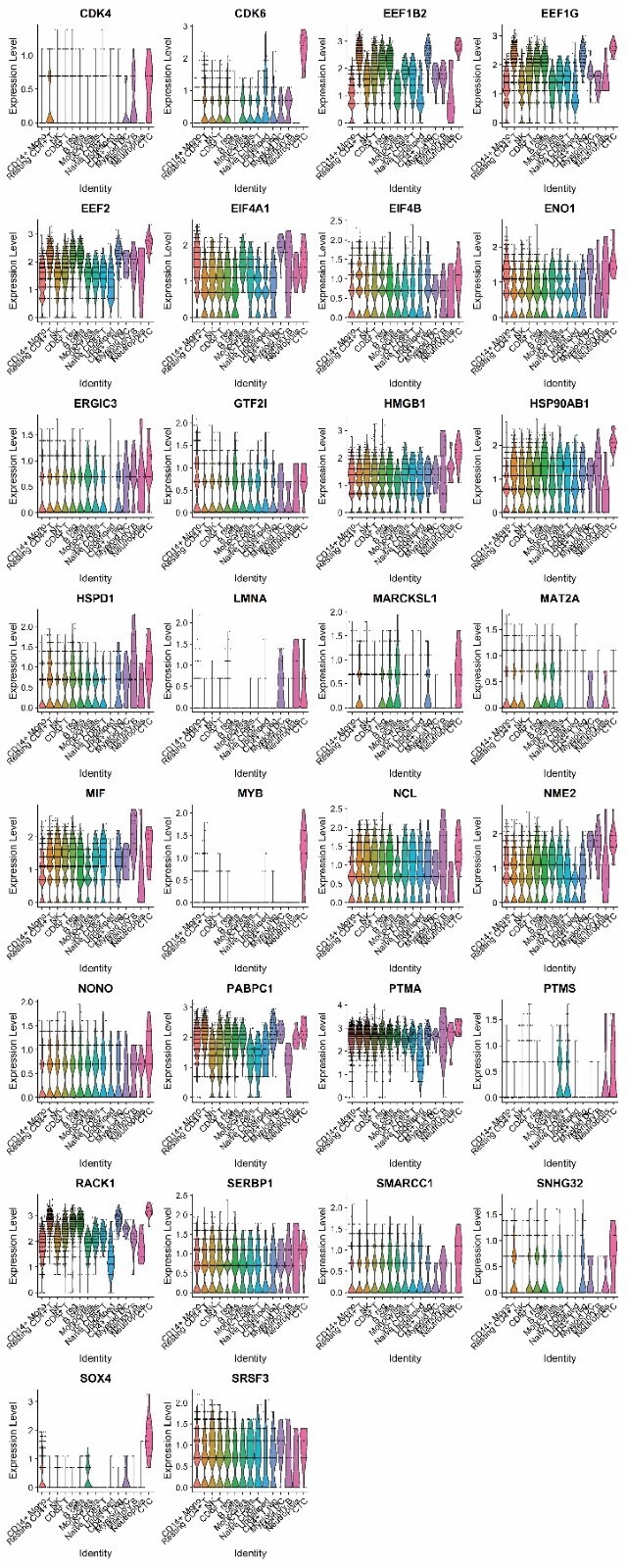

**Supplementary Figure 5.**

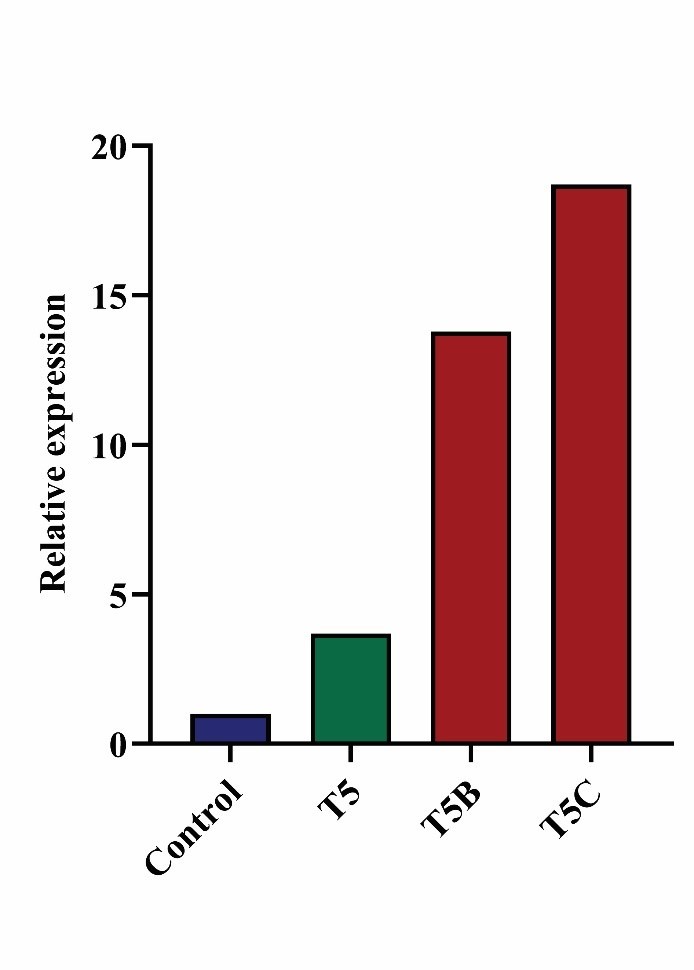

**Supplementary Figure 6.**

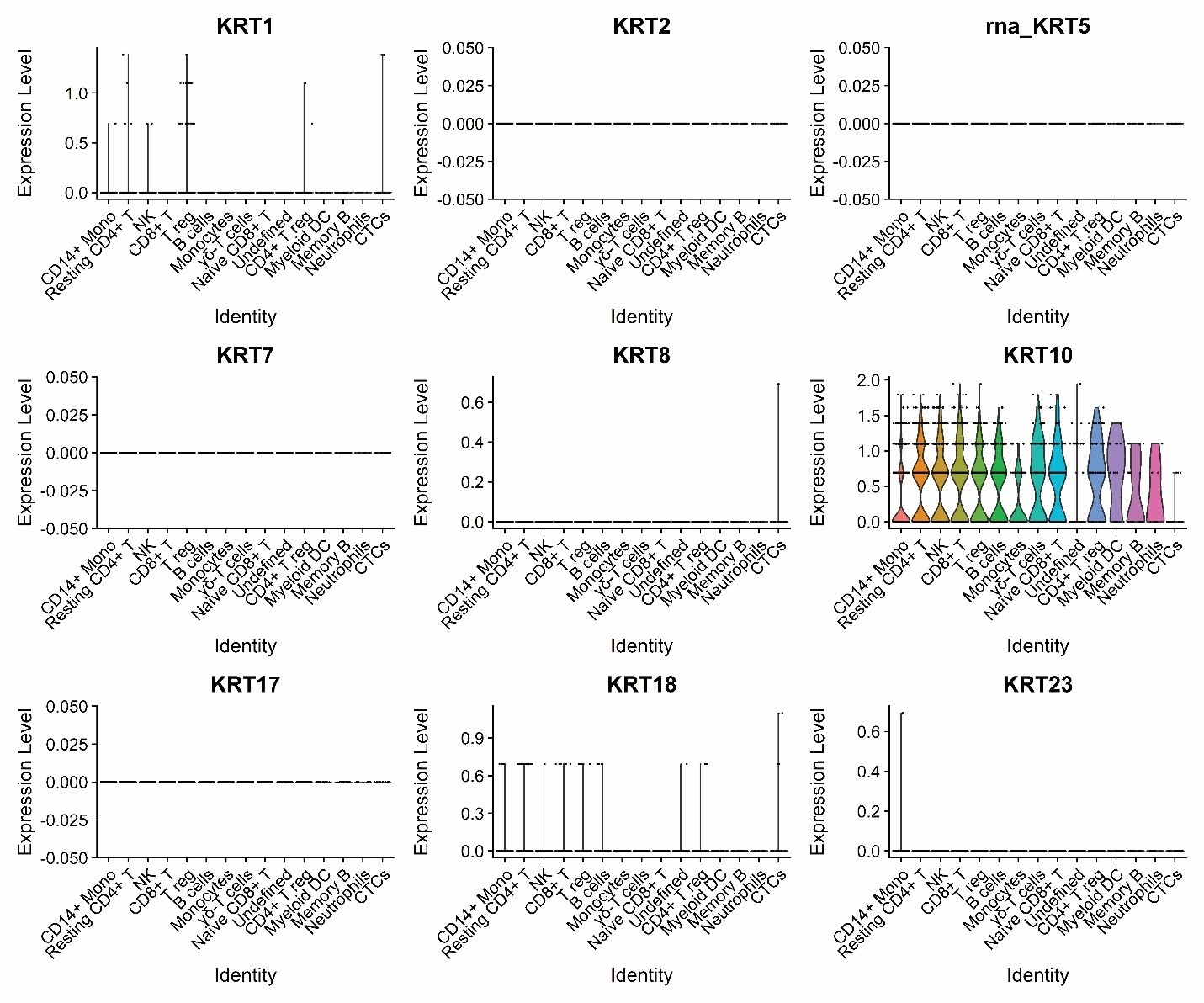

**Supplementary Figure 7.**

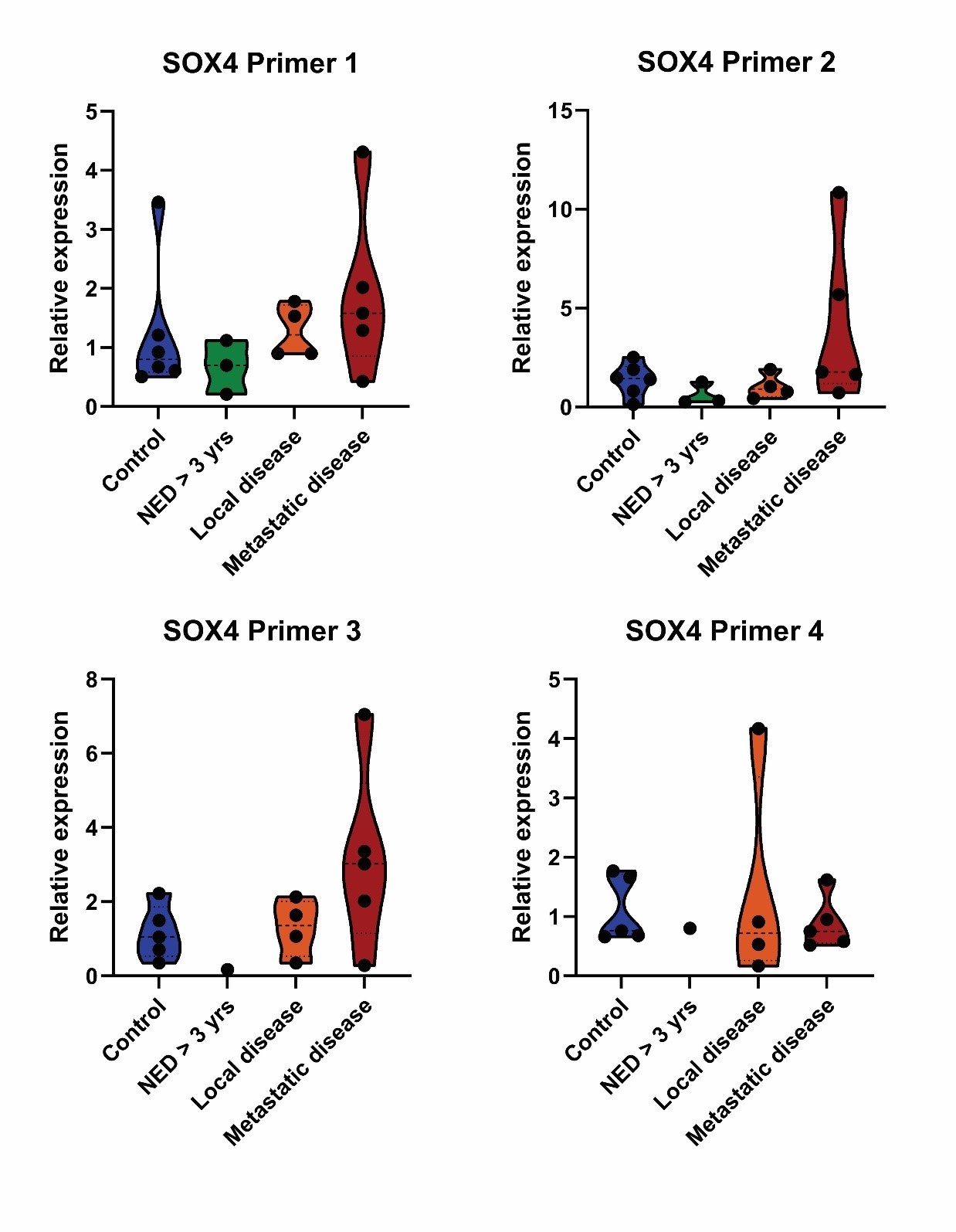

**Supplementary Figure 8.**
