## Supplemental Methods, giving more in depth information on the material for "Circulating Adenoid Cystic Carcinoma associated MYB transcripts enable rapid and sensitive detection of metastatic disease in blood liquid biopsies"

**DETAILED METHODS**

Peripheral blood sample collection

Control samples (n = 23) were collected from healthy individuals with no history of cancer. No evidence of disease (NED) (n =15) samples were obtained from patients with a previous history of ACC but whom had been deemed disease-free for longer than 3yrs at the time of blood collection by current medical standards. Local disease samples (n = 6) were from patients with newly diagnosed and/or locoregionally restricted disease. Metastatic disease samples (n =5) were collected from patients known to have metastatic disease in the lymph nodes, lungs, and liver at the time of blood collection. Samples from patients with multiple blood draws will be labeled by their respective group and patient number, followed by a letter (B, C, etc.) representing the sequential blood draws.

RNA Isolation and cDNA Preparation

Blood samples in EDTA tubes were thawed on ice for 2 hours. RNA was extracted using the New England BioLabs Monarch Total RNA Miniprep Kit (New England BioLabs, Ipswich, MA, USA) according to the manufacturer’s whole blood RNA extraction protocol. Briefly, 200 µL aliquots of a total of 1.2 mL of whole blood were mixed with 200 µL of protection reagent and 10 µL of proteinase K. After a 30-minute incubation, 400 µL of isopropanol was added to each tube, the 800 µL solution was added to a column and spun at 16,000 RPM for 30 seconds. Each tube of 800 µL solution was added to the column until all the sample had passed through. The columns were washed with wash buffer and DNase treatment was added for 15 minutes. Columns were washed priming and wash buffer and RNA was eluted in 50 µL of RNase-free water. For plasma and peripheral blood mononuclear cell (PBMC) RNA extraction, fresh blood samples were spun at 1,900 xG to separate the layers. Each layer was stored in separate cryovial tubes in -80°C freezer until RNA extraction. RNA extraction protocol was the same as whole blood, 600 µL of plasma and ~400 µL of PBMCs were used for extractions.

RNA was quantified and 500-1000 ng of cDNA was made using Thermo Fisher Scientific SuperScript IV VILO Master Mix according to manufacturer instructions (Thermo Fisher Scientific, Waltham, MA, USA). Plasma RNA was not quantified due to low quantity, 10 µL of plasma RNA was used to make cDNA.

Real-time polymerase chain reaction (RT-PCR), Relative expression, and Statistical Analysis

Eleven primer pairs spanning different exons of the MYB gene were tested on preliminary samples (Supplementary Table 1). Primer pairs for the two housekeeping genes RPS18 and UBE2D2 were used for normalization (Supplementary Table 1) (1). RT-qPCR was conducted using the Fast SYBR Green Master Mix (Applied Biosystems, Waltham, MA, USA) on a QuantStudio 7 real-time PCR system (ThermoFisher Scientific). Each reaction included 20 ng of cDNA, 5 µL of master mix, and 1 µL of 10 mM primer forward and reverse primer mix. The RT-qPCR was set up in triplicate with the following parameters: three stages, the hold stage set at 95° C for 20 seconds for 1 cycle, the PCR stage was 95° C for 1 second followed by 60° C for 20 seconds run for 40 cycles, the melt curve stage was 1 cycle of 95° C for 15 seconds, 60° C for 1 minute and 95° C for 15 seconds.

Relative expression of the MYB segment was quantified using the geometric mean of the Ct values for the two housekeeping genes and MYB probe amplification in the control samples (2). Relative expressions were plotted using GraphPad Prism (version 9.3.1). Significance was calculated using a one-way analysis of variance (ANOVA) test with p<0.05 deemed significant. Receiver operating characteristic (ROC) curve was calculated by the Wilson/Brown method and a 95% confidence interval and visualized using GraphPad Prism.

Preparation of peripheral blood mononuclear cell (PBMC) fraction for 5’-capture single cell RNA sequencing (sc-RNAseq)

Blood sample used for sc-RNAseq experiment was collected as described above and prepared using 10x Genomics PMBC preparation protocol. Briefly, PMBCs were separated by centrifugation, followed by red cell lysis (ACK Lysing Buffer, Quality Biological, Gaithersburg, MD), and washed with PBS. The cell pellet was resuspended in 200µL of 1:1 ratio PBS and 0.04% BSA + PBS.

Single-cell RNA sequencing (scRNA-seq) and analysis

Sc-RNAseq was conducted at the Onco-Genomics Core at the University of Miami Miller School of Medicine. Sc-RNAseq library was generated using the 10X Genomics Chromium Single Cell 5’ v2.0 chemistry (10x Genomics, Pleasanton, CA) according to manufactures instructions. The sample was sequenced using a 10B-100 flow cell on an Illumina NovaSeq X (Illumina, San Diego, CA).

CellRanger (10x Genomics Cell Ranger Version 7.1.0) pipeline was used to process and align FASTQ files to the human reference genome GRCh38. Following CellRanger processing, filtering, normalization, PCA, clustering and further downstream analysis was conducted using Seurat R Program (Version 4.3.0) (3, 4). Doublets were removed and cells that had a feature count < 200 and cells with mitochondrial genes > 10% were filtered out. Clustering was performed using FindNeighbors (dimensions = 1:30) and FindClusters (resolution = 0.5). Cell type identities were determined using The Human Protein Atlas, Satija Lab, and previous single cell literature (3-8). Integration of scRNA-seq with previously published spatial transcriptomic sample conducted using Seurat R Program (Version 4.3.0) using FindTransforAnchors.

Supplemental methods references:
